# Role of Intraperitoneal Intraoperative Chemotherapy with Paclitaxel in the Surgical Treatment of Peritoneal Carcinomatosis from Ovarian Cancer. Hyperthermia versus Normothermia. A randomized controlled trial

**DOI:** 10.1101/2021.10.26.21264232

**Authors:** Angela Casado-Adam, Lidia Rodríguez-Ortiz, Sebastián Rufián-Peña, Francisco Cristobal Muñoz-Casares, Teresa Caro-Cuenca, Rosa Ortega-Salas, María Auxiliadora Fernández-Peralbo, María Dolores Luque de Castro, Juan Manuel Sánchez-Hidalgo, César Hervás-Martínez, Antonio Romero Ruiz, Javier Briceño-Delgado, Alvaro Arjona-Sanchez

## Abstract

**Background:** The treatment of ovarian carcinomatosis with cytoreductive surgery and HIPEC is still controversial. The effect and pharmacokinetics of the chemotherapeutics used (specially taxanes) are under consideration at present.

**Methods:** A phase II, simple blind and randomized controlled trial (NTC02739698) was performed. Thirty-two patients with primary or recurrent ovarian carcinomatosis undergoing cytoreductive surgery (CRS) and intraoperative intraperitoneal chemotherapy with paclitaxel (PTX) were included; 16 in hyperthermic and 16 in normothermic conditions. Tissue, serum and plasma samples were taken in every patient before and after intraperitoneal chemotherapy to measure the concentration of PTX. To analyze the inmunohistochemical profile of p53, p27, p21, ki67, PCNA, caspasa-3 and the pathological response a scale of intensity and percentage of expression and a grouped Miller and Payne system were used, respectively. Perioperative characteristics and morbi-mortality were also analyzed.

**Results:** Main characteristics of patients, surgical morbidity, haematoxicity and nephrotoxicity were similar in both groups. The concentration of paclitaxel in the tissue was higher than that observed in plasma and serum, although no statistically significant differences were found between the two groups. No statistically significant association regarding pathological response and apoptosis (caspasa-3) between both groups was proved. Intraperitoneal PTX reduced the expression of p53, p27, p21, ki67 and PCNA more in hyperthermia group, but not significantly.

**Conclusion:** The use of intraperitoneal PTX has proven an adequate pharmacokinetics with reduction of cell cycle and proliferation markers globally without finding differences between its administration in hyperthermia versus normothermia conditions.

## INTRODUCTION

Standard treatment of advanced ephitelial ovarian cancer (EOC), either primary or recurrent, is the complete cytoreductive surgery (CRS) with no residual tumor followed by adjuvant chemotherapy based on taxanes and platinum compounds^1,2^.

Due to its natural history, EOC remains to a great extent localized in the abdominal cavity. Intraperitoneal (IP) chemotherapy allows higher concentrations of chemoterapeutics in the peritoneal cavity than systemic chemoterapy because of its direct administration, improving cytoxicity while minimizing systemic adverse effects^3,4^ and the survival of the patients. Nevertheless, this type of chemotherapy has not been fully accepted because it is not free from complications^5^.

To overcome the IP chemotherapy inconveniences, an intraoperative IP chemotherapy under hyperthermia conditions (HIPEC) arised. Its main objective was to treat residual microscopic disease after CRS, before the formation of adhesions through physical (heat) and chemical (chemotherapeutic) methods^6^. The mechanisms by which HIPEC results in increase tumour response to cytostatics, besides the direct effect of heat^7^ “per se” are multiple^8^; potentiates the cytotoxic effect of some chemotherapeutic agents^9-13^ and increases their tissue penetration^14,15^.

There is a great heterogeneity in fundamental aspects of the administration of HIPEC such as the clinical setting in which it is indicated, the definition of optimal surgery (CC1 vs R1), the cytostatic and its dose, the temperature (37-46ºC) or the perfusion time^16^. The reasons why some authors^17-25^ (our group among them) use taxanes for HIPEC in the treatment of ovarian carcinomatosis are its high efficacy observed in the systemic treatment of EOC and its favorable pharmacokinetics after IP administration due to its high molecular weight^26^ and hepatic metabolism. The theory about an increase in the efficacy of taxanes in their IP administration is supported on different clinical^27-30^ and experimental studies^31,32^.

In an update published by Sugarbaker^33^ it was observed that hyperthermia increases the cytotoxic activity of most citostatics. However, this synergy is not clear in taxanes. Contradictory results have been obtained with respect to the interaction of heat with taxanes^34^, even though they are heat stable and hyperthermia seems to increase the intracelular accumulation of these citostatics.

For all the above, the aim of this study was to analyze the effect of intraoperative IP administration of paclitaxel (PTX) under hyperthermia vs normothermia conditions on antitumor activity, proliferation and cell cycle markers and its pharmacokinetics.

## PATIENTS AND METHODS

A phase II, simple blind, randomized controlled trial (RCT), NTC02739698 was performed.

All steps, including selection of patients, sampling and storage were developed according to the guidelines dictated by the World Medical Association Declaration of Helsinki in 2004. The ethical review board of Reina Sofía Hospital (Córdoba, Spain) approved and supervised the clinical study.

### Inclusion criteria

Age ranging between 18–75 years; histopathologic confirmation of peritoneal carcinomatosis from primary or recurrent EOC (stage IIIb-IIIc FIGO); Karnosfsky index >70 o Gynecologic Oncology Group performance status ≤2 and informed consent form filled by all patients.

### Exclusion criteria

Unfulfillment of inclusion criteria; extra-abdominal metastasis or stage IV FIGO; concomitance of other malignant neoplasm; renal, hepatic or cardiovascular dysfunction; intolerance during treatment or and refusal to participate.

### Sample size calculation

Based on an expected 40% of G3 tumor-regression in experimental arm vs a 1% of G3 tumor-regression in control arm with an α error of 0.05 and β error of 0.20, the sample size was 32 (16 patients pre group).

### Treatment

All patients (except one) received a neoadjuvant chemotherapy regime consisting of 4–6 cycles of carbo-taxol. After confirming the stabilization or regression of disease, the patients underwent optimal CRS followed by intraoperative IP chemotherapy with 60 mg/m^2^ PTX per 2 l of 1.5% dextrose at continuous perfusion for 60 minutes. They were randomized in two groups: the experimental arm (H-group) in which the IP chemoteraphy was administered in hyperthermia conditions (41–42ºC) and the control arm (N-group) where this IP chemotherapy was administered in normothermia conditions (36–37ºC). After surgery most patients received adjuvant carbo-taxol chemotherapy to complete the 8 cycles.

### Variables

Main characteristics of patients, situation of ovarian cancer, previous surgical score (PSS)^35^, response to neoadyuvant chemotherapy (complete response: normalization of Ca 125 and disappearance of signs of disease in radiology tests, partial response: decrease of the value of Ca 125 and decrease of signs of disease in radiology tests according to RECIST criteria^36^) and data of surgery such as peritoneal carcinomatosis index (PCI)^37^ or Completeness of Cytoreduction (CC) Score^38^ were noted.

Dindo-Clavien scale^39^ and CTCAE^40^ v 4.0 were used to describe surgical morbidity and haematological and renal toxicities, respectively. Mortality occurring during the postoperative hospital stay or within 30 days since operation was collected prospectively. Grade 3–4 complications and toxicities were considered as major morbidity.

### Sampling and storage

Two types of peritoneal biopsies (with and without tumor) were taken before and after IP chemotherapy (PRE-chemo and POST-chemo, respectively). Those who have a small and well-perfused area of infiltrated peritoneum were sent to Pathology Department in fresh. The other tumor-free peritoneal biopsies and the blood samples (taken before, immediately after and 1 hour after IP chemotherapy: PRE-chemo, POST-chemo and 1 h POST-chemo, respectively), the latter after centrifugation, were introduced in Eppendorf tubes and preserved at –80ºC to analyze the concentration of PTX in peritoneal tissue by liquid chromatography–tandem mass spectrometry (LC–MS/MS), as reported before^41^.

### Anatomophatologic study

The specimens were fixed in 10% neutral formalin, routinely processed and embedded in paraffin blocks, from which 3 micrometer (μm) thick serial sections were cut and stained with HE, PAS and Masson’s trichome.

The immunohistochemical study was performed using the prediluted antibodies; p53 (clone DO.7. Dako Corporation), p27^kip1^(clone DSC-72. Génova Scientific SL), p21^waf1^(clone polyclonal. Génova Scientific SL), ki67 (clone MIB.1. Dako Corporation), PCNA (clone PC10. Genova Scientific SL) and CASPASA-3 (clone polyclonal. Master Diagnostica SL). For the immunostaining, the Dako EnVision Flex Plus visualization system was used. The sections were examined by two blinded expert pathologists and evaluated by the grouped Miller and Payne (MP) system^42^ for pathological response: G1 (minimal changes that includes MP G1-G2), G3 (microscopic foci, that includes MP G3-G4) and G5 (no residual tumor), according to the percentage of total area involved in the biopsy specimen. Immunohistochemical expression was assessed by percentage of nuclei stained of tumor cells.

### Statistical analysis

Continuous variables were expressed as means and standard deviation (SD), and categorical variables as frequency and percentages. Association between categorical variables was tested with the Pearson’s chi-squared test (χ2). The difference between means of continuous variables was tested by independent *t*-test. A second set of dependent *t*-test (paired-samples *t*-test) was carried out to compare the means of two related groups to determine if a statistically significant difference exists between these means of anatomophatological results. P values ≤ 0.05 were defined as statistically significant.

## RESULTS

### Patients main characteristics

The patient main characteristics are presented in Table 1. No statistically significant differences in preoperative variables were found between the two groups, except BMI, (24.68±2.55 H-group vs 29.00±4.84 N-group, p<0.004).

**Table 1:** Patients main characteristics.

|  | H-group<br>(n=16) | N-group<br>(n=16) | *p* |
| --- | --- | --- | --- |
| Age (years) | 57.06±12.60 | 58.13±9.38 | 0.789 |
| BMI (kg/m<sup>2</sup>) | 24.68±2.55 | 29.00±4.84 | 0.004 |
| Prior abdominal surgery |  |  |  |
| No | 13 (81.2%) | 9 (56.2%) | 0.253 |
| Yes | 3 (18.8%) | 7 (43.8%) |  |
| Prior comorbidity |  |  |  |
| No | 8 (50.0%) | 9 (56.3%) | 1.000 |
| Yes | 8 (50.0%) | 7 (43.8%) |  |
| Ovarian cancer situation |  |  |  |
| Primary | 15 (93.8%) | 12 (75.0%) | 0.330 |
| Recurrent | 1 (6.3%) | 4 (25.0%) |  |
| Prior cancer surgery |  |  |  |
| No | 10 (62.5%) | 4 (25.0%) | 0.075 |
| Yes | 6 (37.5%) | 12 (75.0%) |  |
| Prior Surgical Score (PSS) |  |  |  |
| 0 | 10 (62.5%) | 4 (25.0%) | 0.164 |
| 1 | 4 (25.0%) | 8 (50.0%) |  |
| 2 | 2 (12.5%) | 3 (28.8%) |  |
| 3 | 0 (0.0%) | 1 (6.3%) |  |
| Neoadjuvant chemotherapy |  |  |  |
| No | 1 (6.3%) | 0 (0.0%) | 0.310 |
| Yes | 15 (93.8%) | 16 (100%) |  |
| Response to neoadjuvant chemotherapy |  |  |  |
| Complete | 2 (12.5%) | 4 (25.0%) | 0.714 |
| Partial | 13 (81.2%) | 12 (75.0%) |  |

H-group: Hyperthermia group, N-group: Normothermia group, BMI: Body Mass Index

### Treatment and morbidity

The mean PCI was similar in both groups and although microscopically complete cytoreduction (CC0) was achieved in 81.2% in H-group and 62.5% in N-group, no significant differences were found (Table 2).

**Table 2:** Treatment and morbidity.

|  | H-group<br>(n=16) | N-group<br>(n=16) | *p* |
| --- | --- | --- | --- |
| Ureteral catheterization |  |  |  |
| No | 2 (12.5%) | 1 (6.3%) | 0.544 |
| Yes | 14 (87.5%) | 15 (93.8%) |  |
| PCI | 19.25±6.78 | 21.50±7.81 | 0.391 |
| Peritonectomy procedure |  |  |  |
| Total | 11 (68.8%) | 12 (75.0%) | 0.462 |
| Extensive | 5 (31.3%) | 3 (18.8%) |  |
| Pelvic | 0 (0.0%) | 1 (6.3%) |  |
| CC Score |  |  |  |
| CC0 | 13 (81.2%) | 10 (62.5%) | 0.432 |
| CC1 | 3 (18.8%) | 6 (37.5%) |  |
| Splenectomy |  |  |  |
| No | 12 (75.0%) | 10 (62.5%) | 0.703 |
| Yes | 4 (25.0%) | 6 (37.5%) |  |
| Number of anastomosis |  |  |  |
| 0 | 5 (31.3%) | 8 (50.0%) | 0.363 |
| 1 | 10 (62.5%) | 6 (37.5%) |  |
| 2 | 1 (6.3%) | 2 (12.5%) |  |
| Number of procedures |  |  |  |
| 1-2 | 0 (0.0%) | 1 (6.3%) | 0.665 |
| 3-4 | 11 (68.8%) | 10 (62.5%) |  |
| 5 | 5 (31.3%) | 5 (31.3%) |  |
| Intraoperative blood transfusion |  |  |  |
| No | 5 (31.3%) | 9 (56.3%) | 0.154 |
| Yes | 11 (68.8%) | 7 (43.8%) |  |
| Duration of surgery (minutes) | 492.53±95.81 | 538.06±112.91 | 0.237 |
| Postoperative blood transfusión (units) |  |  |  |
| 0 | 4 (25.0%) | 1 (6.3%) | 0.688 |
| 1-2 | 7 (43.8%) | 7 (43.8%) |  |
| 3-4 | 3 (18.8%) | 5 (31.3%) |  |
| >4 | 2 (12.5%) | 2 (12.5%) |  |
| Postoperative stay (days) | 12.38±6.63 | 13.33±6.65 | 0.691 |
| Surgical Morbidity (Clavien) |  |  |  |
| I-II | 12 (75.0%) | 14 (87.5%) | 0.245 |
| IIIa | 3 (18.8%) | 0 (0.0%) |  |
| IIIb | 1 (6.3%) | 1 (6.3%) |  |
| IVa | 0 (0.0%) | 1 (6.3%) |  |
| IVb | 0 (0.0%) | 0 (0.0%) |  |
| V | 0 (0.0%) | 0 (0.0%) |  |
| Leukopenia (CTCAE 4.0) |  |  |  |
| No | 14 (87.5%) | 15 (93.8%) | 1.000 |
| 1 | 0 (0.0%) | 0 (0%) |  |
| 2 | 0 (0.0%) | 0 (0%) |  |
| 3 | 2 (12.5%) | 1 (6.3%) |  |
| 4 | 0 (0.0%) | 0 (0.0%) |  |
| 5 | 0 (0.0%) | 0 (0.0%) |  |
| Neutropenia (CTCAE 4.0) |  |  |  |
| No | 14 (87.5%) | 15 (93.8%) | 0.491 |
| 1 | 0 (0.0%) | 0 (0.0%) |  |
| 2 | 0 (0.0%) | 0 (0.0%) |  |
| 3 | 1 (6.3%) | 1 (6.3%) |  |
| 4 | 1 (6.3%) | 0 (0.0%) |  |
| 5 | 0 (0.0%) | 0 (0.0%) |  |
| Thombocytopenia (CTCAE 4.0) |  |  |  |
| No | 15 (93.8%) | 13 (81.2%) | 0.478 |
| 1 | 0 (0.0%) | 1 (6.3%) |  |
| 2 | 1 (6.3%) | 2 (12.5%) |  |
| 3 | 0 (0.0%) | 0 (0.0%) |  |
| 4 | 0 (0.0%) | 0 (0.0%) |  |
| 5 | 0 (0.0%) | 0 (0.0%) |  |
| Acute kidney injury (CTCAE 4.0) |  |  |  |
| No | 11 (68.8%) | 6 (37.5%) | 0.346 |
| 1 | 2 (12.5%) | 2 (12.5%) |  |
| 2 | 1 (6.3%) | 4 (25.0%) |  |
| 3 | 2 (12.5%) | 3 (18.8%) |  |
| 4 | 0 (0.0%) | 1 (6.3%) |  |
| 5 | 0 (0.0%) | 0 (0.0%) |  |
| Hematuria (CTCAE 4.0) |  |  |  |
| No | 8 (50.0%) | 7 (43.5%) | 0.931 |
| 1 | 2 (12.5%) | 2 (12.5%) |  |
| 2 | 6 (37.5%) | 7 (43.8%) |  |
| 3 | 0 (0.0%) | 0 (0.0%) |  |
| 4 | 0 (0.0%) | 0 (0.0%) |  |
| 5 | 0 (0.0%) | 0 (0.0%) |  |

There were no treatment-related death. All patients in this study had at least one grade II surgical complication since they all received total parenteral nutrition (TPN) and most of patients required blood transfusion.

Major surgical morbidity (≥ IIIa) was 25% in H-group and 12.5% in N-group. In H-group, two patients had wound infection that needed surgical debridement, one patient had low-grade colorectal fistula treated with conservative treatment and percutaneous drainage and other patient required reintervention due to hemoperitoneum. In N-group, one patient required reoperation due to hemoperitoneum and other patient had septic shock with reintervention and renal and global respiratory failure.

Grade 3–4 hemotoxicity was seen in 18.8% (12.5% H-group vs 6.3% N-group). Grade 3–4 nephrotoxicity was seen in 12.5% of H-group and in 25% of N-group (Table 2).

### Pharmacokinetic

PRE-chemo serum, plasma and tissue samples had PTX values below the detection limit. In H-group the mean PTX concentration in serum POST-chemo and 1 h POST-chemo was 16.61±5.34 ng/ml and 12.18±5.52 ng/ml, respectively, in plasma POST-chemo and 1 h POST-chemo was 17.24±6.14 ng/ml and 11.23±5.15 ng/ml, respectively, and in tissue POST-chemo was 1382±1407.18 ng/ml. In N-group the mean PTX concentration in serum POST-chemo and 1 h POST-chemo was 14.98±4.79 ng/ml and 13.37±4.87 ng/ml, respectively, in plasma POST-chemo and 1 h POST-chemo was 15.14±6.18 ng/ml and 12.36±4.87 ng/ml, respectively and in tissue POST-chemo was 2093.19±1777.92 ng/ml. No significant differences were found between the two groups in any measurement. It was observed that the concentration of PTX obtained at local level (tissue) was much higher than the sistemic (plasma and serum) in both groups (Figure 1).

**Figure 1:**
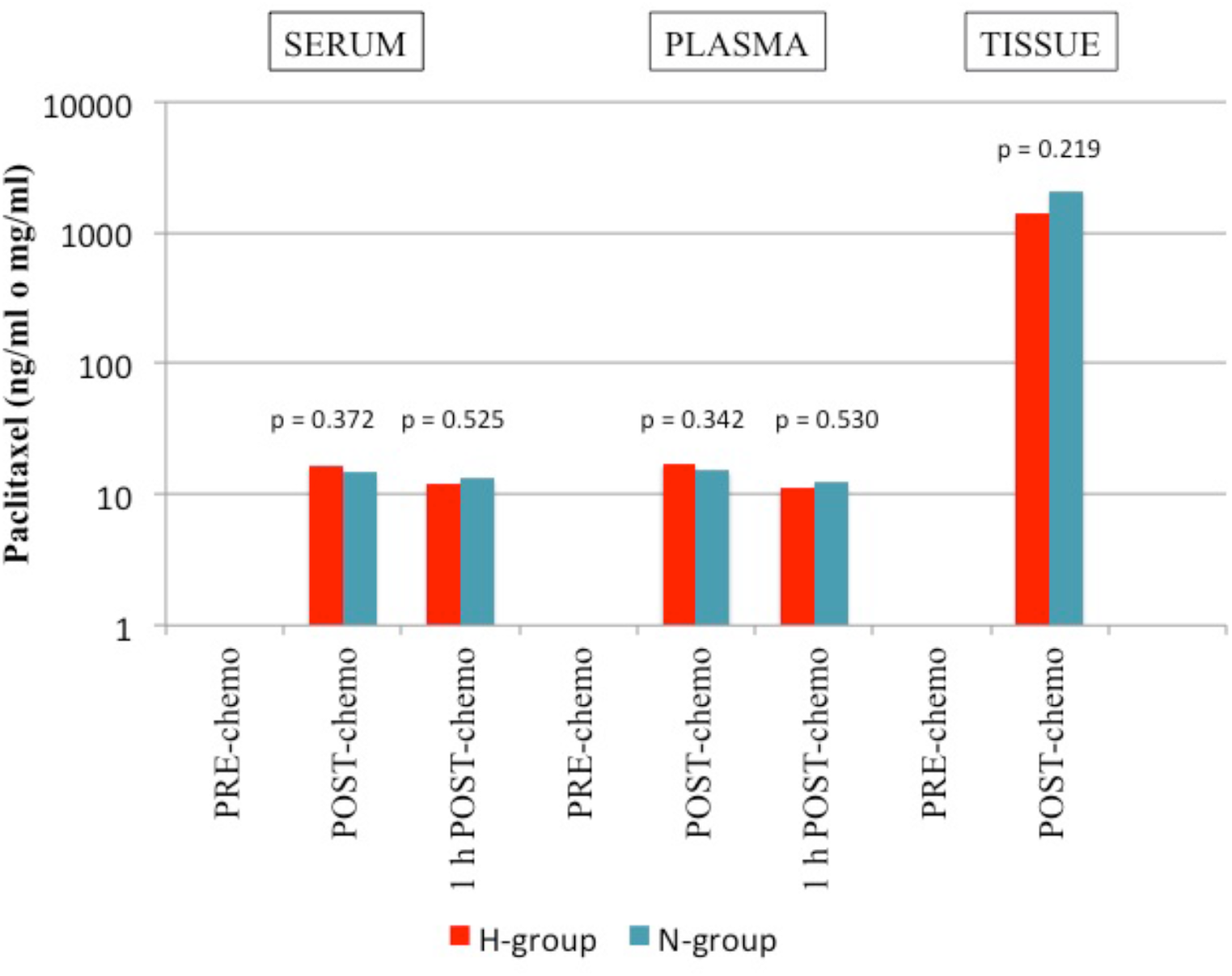
Pharmacokinetics of intraperitoneal PTX admisnistration in our study.

### Anatomophatologic

Regarding to the pathological response according to MP grouped system, no significant differences were observed in both groups. It was observed that in 87.5% of H-group and 81.3% of N-group, IP chemotherapy produced a marked reduction of tumor cellularity. No significant differences in relation to apoptosis (caspase-3) were found either.

The analysis of the results of the cell cycle markers (p53, p27 and p21) shows that there was a significant reduction in the expression of the three markers after IP chemotherapy in the 32 patients (*p*=0.021, *p*=0.000 and *p*=0.000 respectively), but when both groups were compared, this reduction was not statistically significant. Similarly occurred with cell proliferation markers (ki67 and PCNA). After comparing PRE and POST-chemo samples globally, the differences were statistically significant (*p*=0.012 and *p*=0.000), but not when PRE and POST-chemo samples from both groups were compared (Table 3). Although no statistically significant, our results suggest that there is a tendency to decrease the cell cycle and proliferation markers in H-group (Figure 2).

**Table 3:**
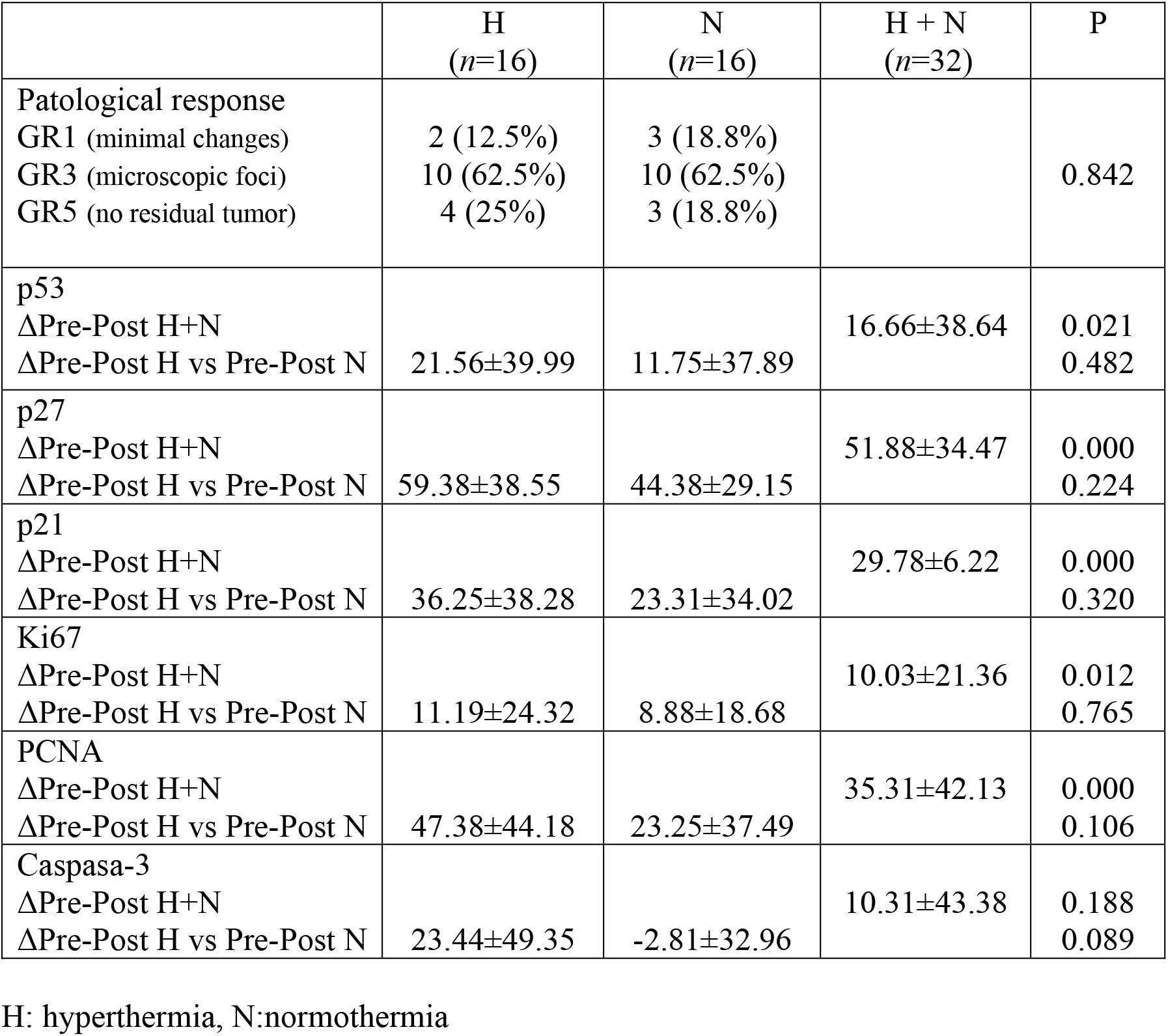
Anatomophatologics results.

|  | H<br>(n=16) | N<br>(n=16) | H + N<br>(n=32) | P |
| --- | --- | --- | --- | --- |
| Patological response |  |  |  |  |
| GR1 (minimal changes) | 2 (12.5%) | 3 (18.8%) |  | 0.842 |
| GR3 (microscopic foci) | 10 (62.5%) | 10 (62.5%) |  |  |
| GR5 (no residual tumor) | 4 (25%) | 3 (18.8%) |  |  |
| p53 |  |  |  |  |
| ΔPre-Post H+N |  |  | 16.66±38.64 | 0.021 |
| ΔPre-Post H vs Pre-Post N | 21.56±39.99 | 11.75±37.89 |  | 0.482 |
| p27 |  |  |  |  |
| ΔPre-Post H+N |  |  | 51.88±34.47 | 0.000 |
| ΔPre-Post H vs Pre-Post N | 59.38±38.55 | 44.38±29.15 |  | 0.224 |
| p21 |  |  |  |  |
| ΔPre-Post H+N |  |  | 29.78±6.22 | 0.000 |
| ΔPre-Post H vs Pre-Post N | 36.25±38.28 | 23.31±34.02 |  | 0.320 |
| Ki67 |  |  |  |  |
| ΔPre-Post H+N |  |  | 10.03±21.36 | 0.012 |
| ΔPre-Post H vs Pre-Post N | 11.19±24.32 | 8.88±18.68 |  | 0.765 |
| PCNA |  |  |  |  |
| ΔPre-Post H+N |  |  | 35.31±42.13 | 0.000 |
| ΔPre-Post H vs Pre-Post N | 47.38±44.18 | 23.25±37.49 |  | 0.106 |
| Caspasa-3 |  |  |  |  |
| ΔPre-Post H+N |  |  | 10.31±43.38 | 0.188 |
| ΔPre-Post H vs Pre-Post N | 23.44±49.35 | -2.81±32.96 |  | 0.089 |

H: hyperthermia, N:normothermia

**Figure 2:**
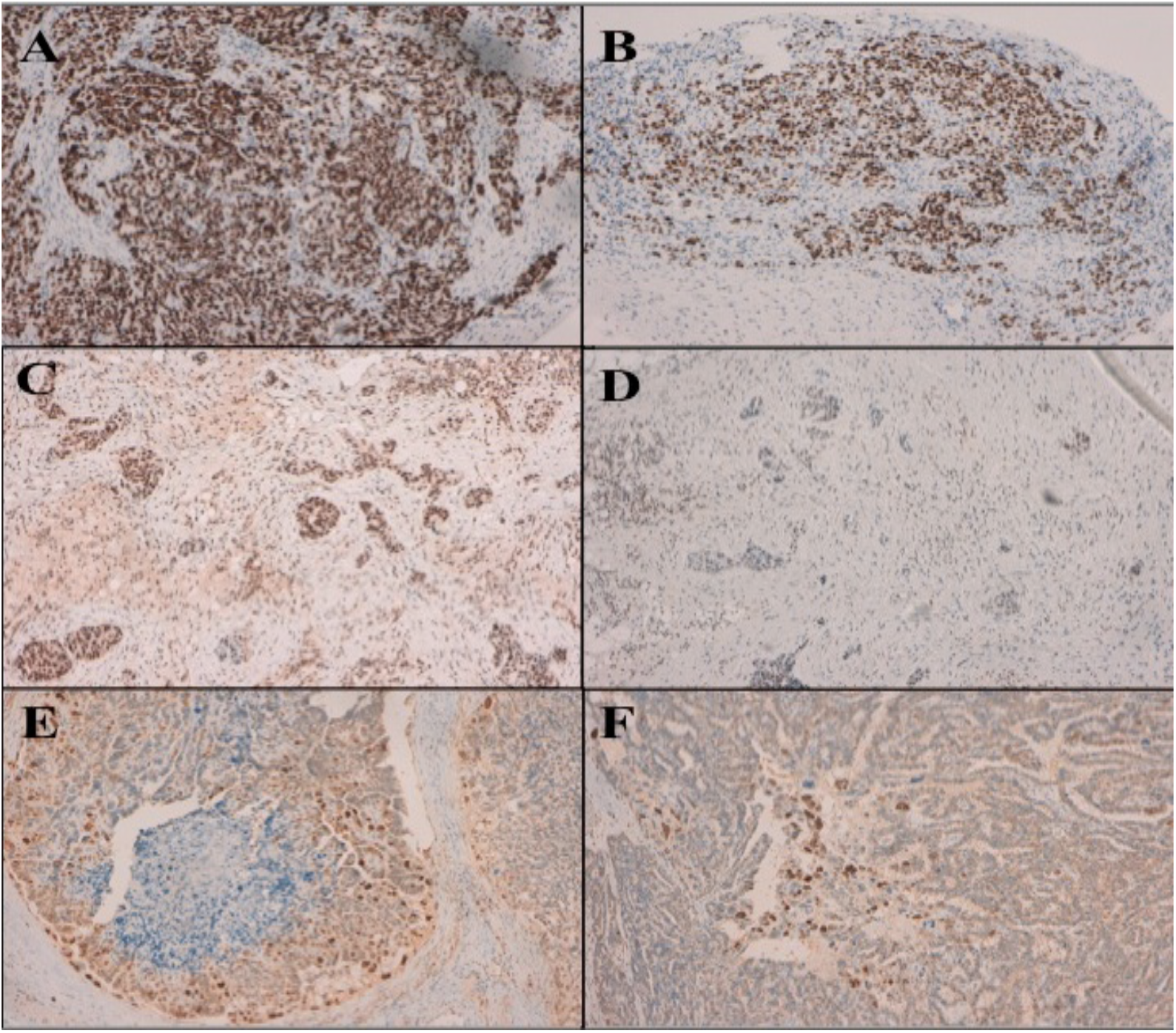
Inmunohistochemical nuclear expression of cell cycle regulatory proteins in 1 H-group patient. A: p53 PRE-chemo, B: p53 POST-chemo, C: p27 PRE-chemo, D: p27 POST-chemo, E: p21 PRE-chemo, F: p21 POST-chemo.

## DISSCUSION

CRS with peritonectomy procedures associated to HIPEC has been worlwide incorporated by numerous medical centers, making this technique the standard of treatment in colon carcinomatosis^43,44^, pseudomyxoma peritonei^45,46^, and mesothelioma^47^. Although the standard treatment of ovarian carcinomatosis is not CRS+HIPEC^1,2^, evidence of its use is growing after recent publications of the first phase III RCT^48^ and a meta-analysis^49^. Spiliotis et al^48^ demonstrated an improvement in survival in the treatment of recurrent EOC with CRS+HIPEC vs CRS alone, where the mean overall survival was 26.7 vs 13.4 months, respectively. The meta-analysis^49^ concludes that CRS+HIPEC improves overall survival rates for both, primary and recurrent EOC vs isolated CRS.

Our group has been carrying out these CRS+HIPEC with PTX in the treatment of ovarian carcinomatosis since 1997^50^. At the beginning it was not always possible to use the perfusion machine that allowed to reach hyperthermia, and IP chemotherapy was administered in normothermia conditions observing how these patients reached similar numbers of survival. This behaviour, added to the contradictory results obtained with respect the interaction of heat with taxanes^33^, led us decide this study.

The analysis of the results shows two homogeneous groups according to pre- and perioperatives variables, except BMI that was significantly higher in N-group. In spite of it, this difference did not affect the ability to achieve optimal CRS that was similar in both groups, which is consistent with the literature as well^51,52^.

The morbidity and mortality outcomes of CRS+HIPEC are similar to a major gastrointestinal surgery, with a surgical morbidity rates ranging from 12% to 52% and a mortality rates ranging from 0.9% to 5.8%^53^. In our experience, the total major surgery morbidity was 25% in H-group and 12.5% in N-group, although twice as much, no significant differences were found between the two groups. These values are within the range of publications of this type of procedure.

Major systemic chemotherapy-related complications as hematological and renal toxicity in CRS + HIPEC ranged from 0% to 28 % and 0% to 18.6%, respectively^53,54^. Hematological and renal major toxicities related to IP PTX administration ranged from 10.5% to 84.2% and 0% to 7%, respectively^5,55^. And for HIPEC PTX administration, major hematological toxicity is reported from 0 % to 13%^20,56^ and renal toxicity above 11,6%^57^. In our study we observed global mayor hematological complications in 12.5% of H-group and 6.3% of N-group and a mayor renal toxicity in 12.5 % of H-group and 25% of N-group.

In our study the maximal tissue concentration was average 84.54 and 178.01 time longer than the maximal plasma concentration (H and N-group, respectively), a fact that supports that PTX is a chemotherapist suitable to IP administration and is in keeping with previous reports of intraperitoneal use of PTX^20,58^. Although PTX concentration in the N-group was almost twice that of H-group, this fact may be related to the contradictory results of the effect of hyperthermia on the pharmacokinetics of taxanes^34^, no significant differences were observed in both groups.

To assess the effect of IP administration of PTX on the pathological response (reduction in tumor cellularity) we have used the grouped Miller and Payne system, widely studied in the effect of neoadyuvancia on locally advanced breast cancer^59^, but not used previously in the treatment of ovarian cancer with HIPEC. In our study, although we did not find significant differences in both groups, it was observed that in 87.5% of H-group and 81.3% of N-group, IP chemotherapy produced a marked reduction of tumor cellularity.

Proliferation and cell cycle control are central processes in the biology of cancer^60^ and our study has shown for the first time the behavior of these biomarkers to IP chemotherapy. As p53 mutation seems to be related to the development of chemoresistance and recurrence^61,62^, findings regarding the predictive and prognostic relevance of expression of p21 and p27 in EOC are inconsistent^63-66^.

In our study we observed a statistically significant difference when comparing the PRE-chemo samples with the POST-chemo of the 32 patients in both cell cycle and proliferation markers; but comparing the differences between the PRE-chemo with the POST-chemo samples of each group, the results were not statistically significant. Although no significant, our results suggest that there is a greater decrease in these tumor markers in H-group: a fact that could be related to the cytotoxic effect “per se” of hyperthermia.

The main limitation of the study is the small sample size, even though our hospital is a reference for the treatment of ovarian carcinomatosis in our country, so it will be necessary additional studies with a larger sample size to validate the impact of the temperature in IP administration of PTX on pharmacokinetics, pathological response and cell cycle markers. Nevertheless, to our knowledge, this is the first study in ovarian carcinomatosis where the IP chemotherapy is compared with taxanes in hyperthermia vs normothermia conditions.

In conclusion, in this clinical trial PTX has proven to have an adequate pharmacokinetics to treat ovarian carcinomatosis, reaching optimal concentrations in tissue and minimal in serum and plasma as well as a reduction of cell cycle and proliferation markers globally when administered in the peritoneal cavity during CRS. Although no significant differences were found between the two groups, it seems that hyperthermia may have a negative effect on the pharmacokinetics of PTX, but could enhance its cytotoxic effect, making this combination an effective treatment.

## Data Availability

All data produced in the present work are contained in the manuscript

## ACKNOWLEDGEMENTS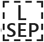

All staff who have participated directly or indirectly in the realization of this study, specially the Department of Analytical Chemistry of the University of Córdoba, the Department of Pathology and the Oncologic and Pancreatic Surgery Unit of Reina Sofia Hospital is thanked.

